# Socio-ecological heterogeneity and uncertainty in the elimination of human schistosomiasis

**DOI:** 10.1101/2024.03.20.24304586

**Authors:** M. Inês Neves, Gregory C Milne, Joanne P. Webster, Martin Walker

## Abstract

Schistosomiasis is a neglected tropical disease caused by parasitic flatworms which infect approximately 240 million people worldwide living in poverty, mostly in sub-Saharan Africa. The World Health Organization (WHO) aims to eliminate schistosomiasis by 2030, predominantly relying on a strategy of mass drug administration (MDA) using praziquantel. The effectiveness of MDA can vary widely among disease foci. Mathematical modelling is increasingly being used to understand the transmission dynamics of schistosomiasis and for predicting the effectiveness of MDA and time frames to elimination. Due to the highly focal nature of schistosomiasis, many key parameters influencing its transmission are likely highly geographically variable. Yet typically models do not fully integrate this uncertainty into predictions. This can lead to unrealistic expectations on the prospects for elimination. Here, we present a schistosomiasis transmission model to evaluate how uncertainty in parameters relating to local socio-ecological conditions influence resilience of the parasite population to intervention and the effectiveness of MDA. We discuss the growing importance of incorporating uncertainty in mathematical models for this and other NTDs to enable transparent communication of predictions as we move towards the 2030 elimination goals.

## 1. Background

Schistosomiasis is targeted by the World Health Organization (WHO) for elimination by 2030, principally using a strategy of preventive chemotherapy by mass drug administration (MDA) with praziquantel [1]. Praziquantel is a safe and efficacious treatment which, if distributed sufficiently frequently and at adequate population coverage, is thought to be compatible with reaching elimination in low to moderate prevalence settings [2-5]. Yet increasingly, areas of persistent infection—often termed ‘hotspots’ or ‘biological hotspots’—have been documented that cannot be explained by inadequate treatment frequency, coverage or other operational programmatic shortcomings [6-13].

In 2022, the WHO released a provisional definition of schistosomiasis hotspots as “communities with prevalence of *Schistosoma spp*. infection ≥ 10% that demonstrate lack of an appropriate response to two annual rounds of preventive chemotherapy, despite adequate treatment coverage ≥ 75%” [14]. More generally, the term embodies the epidemiological circumstance where the prevalence of infection or—less commonly measured—the intensity of infection, remains stubbornly high despite well-implemented interventions [7,8,13]. Heterogeneity in pre-intervention transmission conditions— reflected in baseline measures of prevalence and intensity (the only epidemiological data routinely measured by MDA programmes)—are a natural explanation for this phenomenon [15]. However, multi-country epidemiological data collected by the Schistosomiasis Consortium for Operational Research and Evaluation (SCORE) [16] found that hotspots could not be solely and consistently explained by baseline prevalence indicators or programmatic deficiencies in the delivery of MDA [10, 17]. Rather, a variety of meteorological and ecological factors were also associated with ‘hotspot’ communities [18]. Furthermore, population genetic studies have also indicated further biological fitness traits inherent within certain parasite populations that may also been associated with ‘hotspots’ [13, 19]. Accordingly, particularly with the inclusion of persistent ‘hotspots’ as a prescription measure identifying the type of treatment regime to be initiated in a region, subsequent studies have proposed the need for a revision of this term, encompassing at least geographic focality, as well as measures of both prevalence and intensity [20].

Mathematical modelling has been used for more than half a century to understand the population biology and transmission dynamics of schistosomes [21-24] and more recently for predicting the effectiveness of MDA programmes [2-5]. Typically, models are parameterized to different endemic settings by varying the basic reproduction number, *R*_0_. This defines the intensity of transmission (force of infection) and associated resilience of the parasite population to intervention; the higher the *R*_0_, the higher the force of infection and the greater the intervention effort required to achieve elimination. Most models do not consider uncertainty in the parameters that define *R*_0_ (but see [25, 26]), resulting in each *R*_0_ value being uniquely determined by a particular parameter combination (by contrast, stochastic uncertainty is well covered by existing models, e.g., [5, 10, 27, 268]). Yet, different combinations of (uncertain) parameters may yield the same *R*_0_ albeit inducing different model dynamics under intervention.

While uncertain, several model parameters that capture fundamental aspects of schistosome population biology, such as worm and snail mortality rates [23,24], are likely to be geographically relatively more homogenous relative to those that relate to local human socio-ecological conditions. These include parameters that capture human contamination of water bodies with schistosome eggs via stool or urine (influenced by, for example, access to water, sanitation and hygiene [29], human population density and local behaviour [30]) and exposure to infectious cercariae (driven by, for example, water contact patterns [31], snail density and differences in water temperatures that may affect the rate of cercarial shedding and viability by infectious snails [32]). It is therefore likely that local heterogeneity in the drivers of transmission may engender different infection dynamics during interventions, despite similar baseline epidemiological indicators (prevalence/intensity). Certain parameters may also be particularly important in determining time frames to specific ‘elimination’ endpoints, such as elimination as a public health problem (EPHP, defined as 1% prevalence of heavy infections one year after the previous MDA round) and interruption of transmission (IoT, i.e., suppressing the parasite load below the breakpoint such that the population goes into terminal decline) [1], and could thus provide a target for complementary strategies to accelerate time frames to elimination.

In this study, we use a simple schistosomiasis transmission model to explore how heterogeneity in local socio-ecological conditions affects the transmission dynamics of schistosomiasis and its resilience to intervention. We identify specific sources of variation that make schistosomiasis more difficult to eliminate, results that have important implications for modelling the impact of interventions and for designing effective complementary strategies to enhance the likelihood of reaching elimination. More broadly, this study highlights the importance of incorporating parametric uncertainty in mathematical transmissions models for open and transparent communication of modelling results to stakeholders and for managing expectations on the precision of model predictions.

## 2. Materials and Methods

### (a) Schistosomiasis transmission dynamics model

The model is a simplified and compressed version of that originally presented by Anderson & May [23,24]. A complete derivation of the model is given in the Supplementary Information S1. Parameter and function definitions are given in Table 1. Briefly, the basic reproduction number, *R*_0_ = *R*_*H*→*S*_*R*_*S*→*H*_—defined as the average number of female schistosomes produced by a single fertile female schistosome in the absence of density-dependent effects—is comprised of two components, that from humans to snails 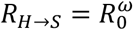, and from snails to humans, 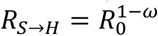. Parameter *ω* ∈ (0,1), describes asymmetry in human-to-snail versus snail-to-human transmission such that as *ω* → 1, transmission is dominated by human-to-snail transmission (i.e., contamination) and vice versa for *ω* → 0 (i.e., human exposure).

**Table 1.**
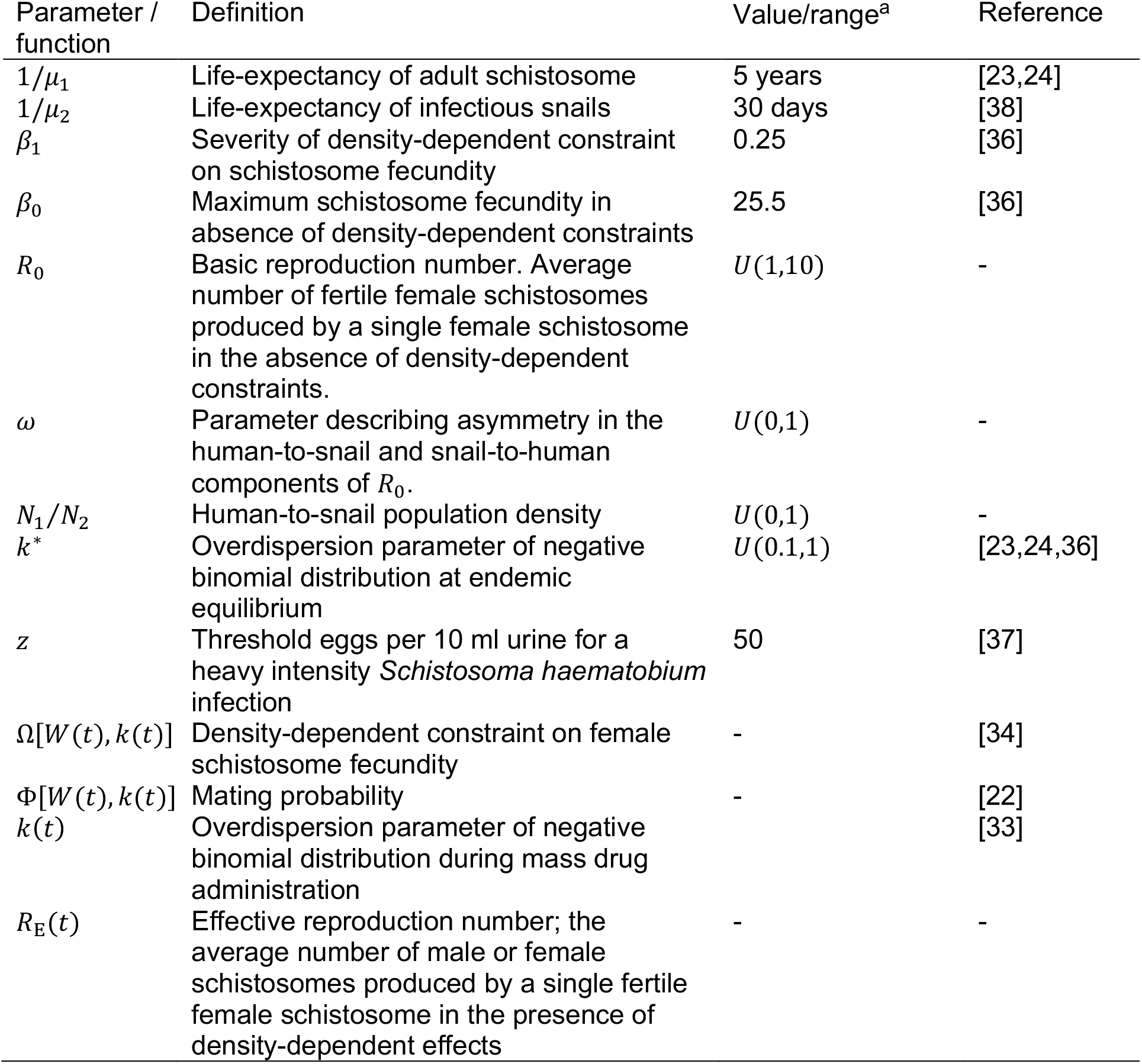
Model parameter definitions, values and functions.

By assuming that the dynamics of infectious snails occurs on a much shorter time scale than that of adult schistosomes, the proportion of infectious snails can be set to its equilibrium position, *y*^*^, with respect to the mean number of adult schistosomes per human, *W*(*t*). This allows derivation of a single ordinary differential equation (ODE) that describes the dynamics of infection,

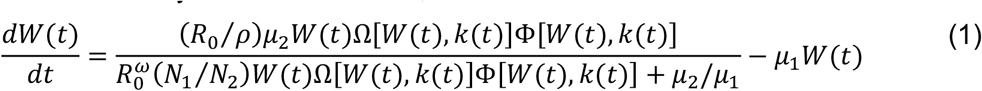

Here, *μ*_1_ and *μ*_2_ are the per capita mortality rates of adult schistosomes and infectious snails respectively, *ρ* is the sex ratio (proportion female), *N*_1_/*N*_2_ is the human-to-snail population density, *k*(*t*) is the overdispersion parameter of a negative binomial distribution that (inversely) describes the degree of schistosome aggregation among hosts. The time dependency in *k*(*t*) results from the effects of variable adherence patterns during MDA on the distribution of schistosomes among hosts, which remains adequately approximated by a negative binomial distribution [33]. The functions Ω(·) and Φ(·) capture density-dependent effects on transmission imposed by density-dependent fecundity [34] (parameterized for *S. haematobium* [35,36]) and the mating probability [22] respectively (see Supplementary Information S2 for mathematical definitions of Ω(·), Φ(·) and *k*(·)).

### (b) Stable/endemic and breakpoint/unstable equilibria

The equilibria of the model are found by setting the derivative on the left-hand side of Equation (1) to zero and rearranging such that,

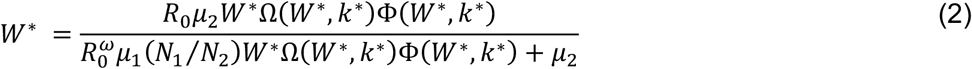

There exist three solutions to Equation (2) corresponding to the trivial equilibrium (*W*^*^ = 0), the unstable/breakpoint equilibrium—below which the parasite population cannot sustain itself—and the endemic/stable equilibrium [23,24]. Note that *k*^*^ is an input parameter to the model, corresponding to the overdispersion of parasite among hosts at stable/endemic equilibrium. Hence, unstable/breakpoint values of *W*^*^ implied by Equation (2) are conditional on a static value of *k*^*^and do not account for the dynamic *k*(*t*) induced by MDA [33].

### (c) Effective reproduction number

The effective reproduction number is defined as the average number of male or female schistosomes produced by a single fertile female schistosome. Therefore, when *R*_E_(*t*) < 1, the parasite population cannot replenish itself and declines terminally to zero [34]. *R*_E_ (*t*) is given by

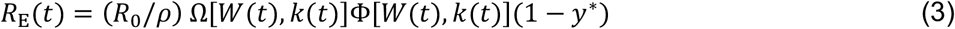

where (1 − *y*^*^) is the proportion of susceptible (uninfected) snails in a population. Note that unlike the unstable/breakpoint *W*^*^ from Equation (2), the criteria *R*_E_ (*t*) < 1 accounts for the dynamics in *k*(*t*) induced by MDA [33] and is therefore useful for determining timescales to elimination of transmission under MDA intervention.

### (d) Prevalence and intensity of infection

We parameterized the model for *S. haematobium* using the density-dependent relationship between egg output (eggs per 10 ml of urine; infection intensity), *E*(*t*), and worm burden described in Neves et al. [35,36],

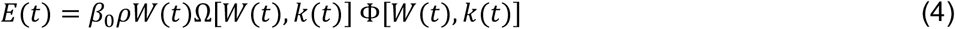

where *β*_0_ is the maximum fecundity of a female schistosome in the absence of density-dependent constraints. Again, invoking the assumption of a negative binomial distribution of schistosomes among hosts, the prevalence of eggs *p*(*t*) (i.e., the prevalence of mated female worms releasing eggs), is given by

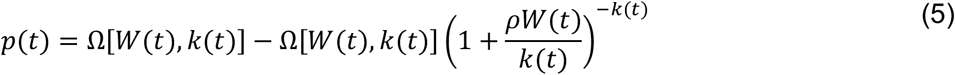

and the prevalence of heavy infection, *p*_*H*_(*t*), is given by

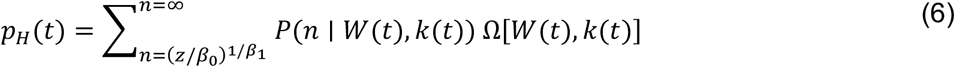

where *z* denotes the 50 eggs per 10 ml urine threshold for heavy *S. haematobium* infection used by the WHO [37]. Note that unlike *p*(*t*), the expression for *p*_*H*_(*t*) has no convenient closed form (see Supplementary Information S3 for details) and so was calculated numerically at every time step of the model.

### (e) Socio-ecological heterogeneity

The transmission model parameters in Table 1 were divided into those that are likely to vary geographically across transmission foci due to differences in local socio-ecological conditions, versus those for which geographical heterogeneity is likely less pronounced. We propose that asymmetry in the components of the basic reproduction number, defined by *ω*, will likely vary according to local socio-economic (e.g. access to water, sanitation and hygiene [29]), behavioural (e.g., water contact patterns [31]) and environmental (e.g. differences in water temperatures that may affect the rate of cercarial shedding by infectious snails [32]) conditions. Similarly, we propose also that *N*_1_/*N*_2_ and *k*^*^ will be highly geographically variable (the latter, for example, may be driven by local individual exposure patterns [31]). To capture this parametric variability, we sampled 5,000 parameter sets of *R*_0_, *ω, N*_1_/*N*_2_ and *k*^*^ from uniform distributions representing a spectrum of plausible transmission conditions (Table 1) using a Latin hypercube to ensure representative coverage of the four-dimensional parameter space.

### (f) Simulating mass drug administration

We simulated mass drug administration by assuming that a fraction of adult schistosomes—given by the product of the treatment coverage and praziquantel efficacy—are killed instantaneously at the same point each year (see [2,40] for a similar example of this approach). Treatment coverage was assumed to be 50% (to crudely account for treatment being targeted predominantly at school-aged children) and the efficacy of praziquantel was assumed to be 94% [39].

### (e) Elimination criteria

Two endpoints for schistosomiasis elimination were chosen based on the 2030 WHO roadmap goals for schistosomiasis elimination [1]. Elimination as a public health problem (EPHP) is defined as reaching less than 1% prevalence of heavy-intensity infections (<50 eggs per 10 ml of urine for *S. haematobium*); interruption of transmission (IoT) is defined by the parasite population tending terminally to zero (i.e., the population having been suppressed below the breakpoint, *R*_E_(*t*) < 1) [22-24,34,39]. The number of MDA rounds required to reach EPHP and IoT for each parameter set from the LHS was determined by simulating 25 annual MDA rounds and checking one time step before the subsequent treatment round whether each criterion had been reached.

### (f) Partial rank correlation coefficients

Partial rank correlation coefficients (PRCCs) were used to quantify the association between the (variable) parameters *R*_0_, ω, *N*_1_/*N*_2_ and *k* and the stable and unstable equilibria of *E*(*t*) and the number of MDA rounds to reach EPHP or IoT. See Supplementary Information S5 for details.

## 3. Results

### (a) Socio-ecological heterogeneity and endemic transmission

The basic reproduction number *R*_0_ is composed of two components, transmission from human to snails, *R*_*H*→*S*_, and from snails to humans, *R*_*S*→*H*_. Transmission can be symmetrical (*ω* = 0.5) or dominated by either host contamination (*ω* → 1) or exposure (*ω* → 0) (Figure 1a). The stable/endemic mean infection intensity per host, *E*^*^ (here parameterized for *S. haematobium*, i.e., eggs per 10 ml of urine), shows a noisy and non-linear relationship with *R*_0_ (Figure 1b, purple points) due to the influence of other heterogenous variables. For a given *R*_0_, *E*^*^ decreases (negatively correlated) with increasing *ω* (i.e., asymmetry towards human-to-snail transmission), increasing *k*^*^ (i.e., *decreasing* parasite overdispersion) and increasing *N*_1_/*N*_2_ (i.e., decreasing density of snail intermediate hosts; Figure 1c). There exists a similar noisy and non-linear relationship between stable/endemic egg prevalence, *p*^*^ and *R*_0_ with similar effects of *ω* and *N*_1_/*N*_2_, albeit with an opposite correlation with *k*^*^; *p*^*^ is positively correlated with increasing *k*^*^ (i.e., decreasing parasite overdispersion) because fewer hosts are uninfected when parasite overdispersion declines.

**Figure 1.**
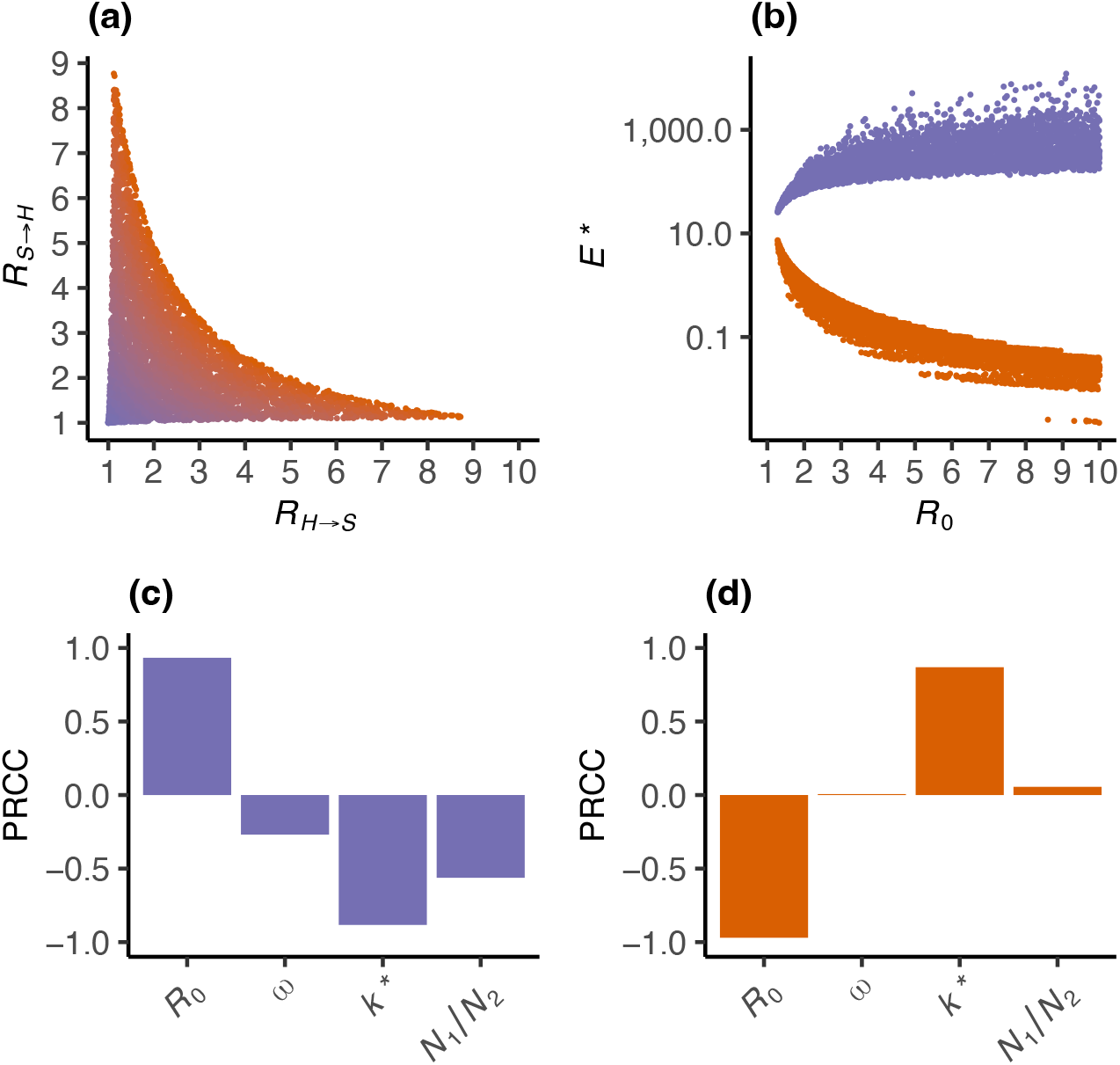
Influence of socio-ecological heterogeneity on stable/endemic and unstable/breakpoint *Schistosoma haematobium* infection intensity. Variation in the stable/endemic (purple points and bars) and unstable/breakpoint (red points and bars) equilibrium mean infection intensity per host, *E*^*^ (eggs per 10 ml of urine), is generated using 5,000 parameter sets from a four dimensional Latin hypercube sample of the basic reproduction number, *R*_0_, parameter *ω*, defining asymmetry in the human-to-human and snail-to-human components of *R*_0_, *R*_*H*→*S*_ and *R*_*S*→*H*_, the overdispersion of schistosomes among hosts at endemic stability, *k*^*^, and the human-to-snail population density, *N*_1_/*N*_)_. Panel (a) shows the relationship between *R*_*H*→*S*_ and *R*_*S*→*H*_ arising from sampling *ω* ∈ (0,1) and *R*_0_ ∈ (1,10), with colours from purple to red indicating increasing *R*_0_. Panel (b) shows the relationship between the stable/endemic (purple) and unstable/breakpoint (red) *E*^*^ and *R*_0_, with variation driven by parameters *ω, k*^*^ and *N*_1_/*N*_2_. Panels (c) and (d) show the influence of *R*_0_, *ω, k*^*^ and *N*_1_/*N*_2_ on the stable/endemic *E*^*^ (purple bars) and the unstable/breakpoint *E*^*^ (red bars) using partial rank correlation coefficients (PRCCs). The PRCC quantifies the correlation between each variable and *E*^*^, while controlling for the effects of other variables.

### (b) Socio-ecological heterogeneity and unstable breakpoints

The relationship between *R*_0_ and the unstable/breakpoint mean infection intensity per host, *E*^*^, is shown in Figure 1b (red points). The unstable breakpoint is strongly negatively correlated with *R*_0_ (i.e., larger *R*_0_ yields a smaller breakpoint, such that IoT is more difficult to achieve) and positively correlated with increasing *k*^*^ (i.e., decreasing overdispersion leads to a larger breakpoint, such that IoT is easier to achieve). Unlike the stable/endemic *E*^*^, *ω* and *N*_1_/*N*_2_ have a negligible impact on the unstable breakpoint (i.e., PRCC values ≈ 0; Figure 1d). Similar effects of *R*_0_, *ω, k*^*^ and *N*_1_/*N*_2_ are apparent on the unstable/breakpoint egg prevalence, *p*^*^(Supplementary Information Figure S1).

### (c) Treatment dynamics

The dynamics in infection intensity, *E*(*t*), egg prevalence, *p*(*t*), overdisperion, *k*(*t*) and the effective reproduction number, *R*_E_ (*t*) are illustrated for five annual rounds of MDA (50% coverage) in Figure 2. Even for similar stable/endemic mean infection intensity per host, *E*^*^, there is considerable variation in the dynamics, *E*(*t*) (Figure 2a) and *p*(*t*) (Figure 2b; the latter driven largely by heterogeneity in *k*^*^ and the ensuing dynamics in *k*(*t*) Figure 2c) and whether the outcome of MDA is interruption of transmission (IoT; *R*_E_(*t*) < 1) or resurgence (Figure 2d). The infection intensity at which IoT is reached is approximately linearly related to the unstable/breakpoint intensity (Figure 3a) but for prevalence the relationship is highly non-linear, with the prevalence at which IoT is reached generally much lower than the corresponding unstable/breakpoint prevalence (Figure 3b).

**Figure 2.**
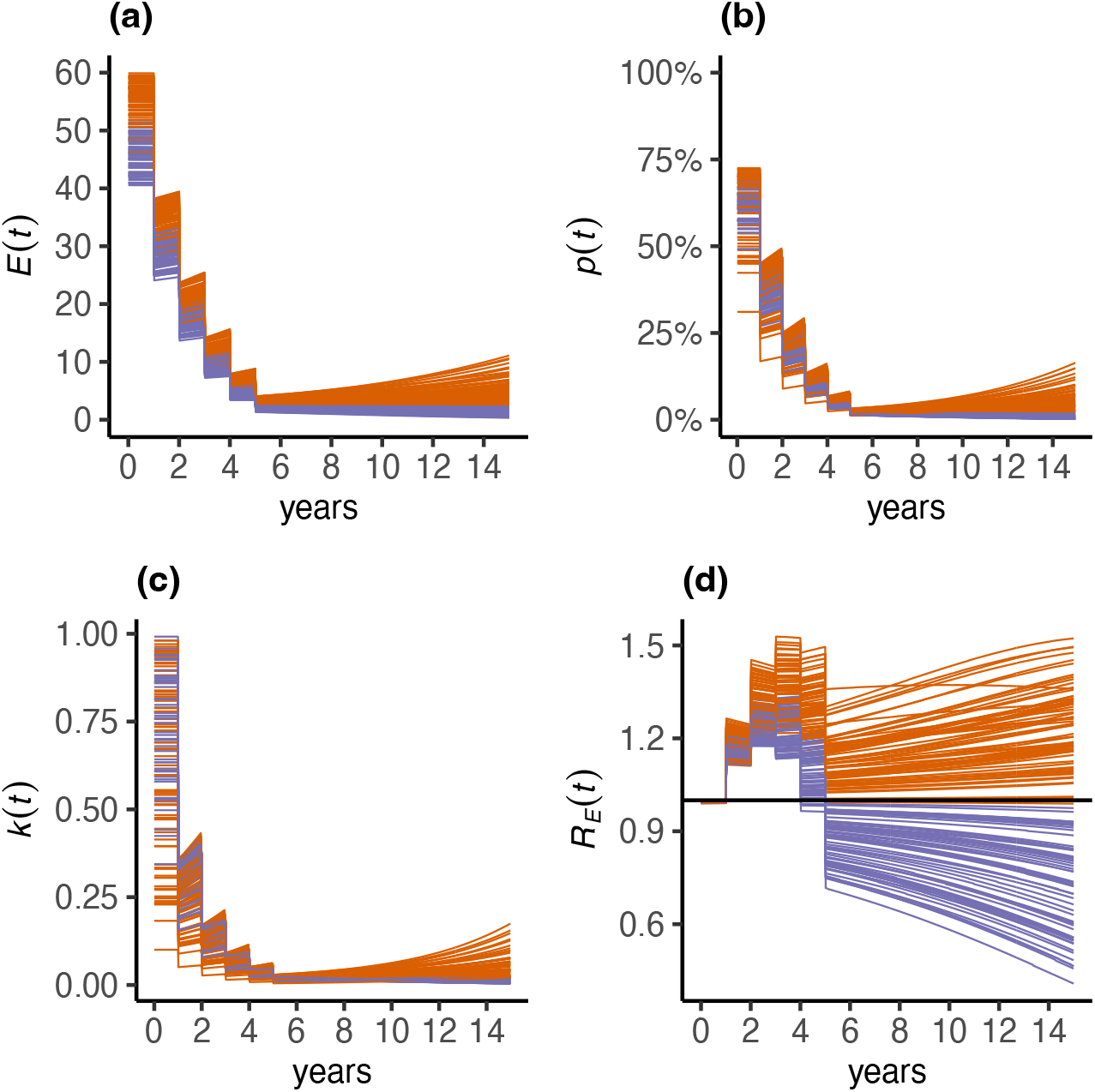
Influence of socio-ecological heterogeneity on urogenital schistosomiasis dynamics through mass drug administration. The mathematical transmission model was used to simulate infection dynamics during five annual rounds of mass drug administration (MDA) with praziquantel assuming a coverage of 50% and drug efficacy of 94%. Variation in transmission dynamics is generated using 5,000 parameter sets from a four-dimensional Latin Hypercube sample of parameter *ω*, defining asymmetry in the human-to-human and snail-to-human components of the basic reproduction number, *R*_*H*→*S*_ and *R*_*S*→*H*_, the overdispersion of schistosomes among hosts at endemic stability, *k*^*^, and the human-to-snail population density, *N*_1_/*N*_2_. Panel (a) depicts the variable dynamics of *Schistosoma haematobium* infection intensity (eggs per 10 ml urine), *E*(*t*), from parameter sets with endemic equilibrium mean infection intensity, *E*^*^, of 50 (± 10) eggs per 10 ml urine. Panel (b), (c) and (d) depict, respectively, the corresponding dynamics for egg prevalence, *p*(*t*), the overdispersion of schistosomes among hosts *k*(*t*) and the effective reproduction number, *R*_E_(*t*). Simulations that result in interruption of transmission (resulting from passing the breakpoint, *R*_E_(*t*) < 1, and leading to a terminal decline in parasite population density towards zero) are coloured purple, and those that resurge (after cessation of MDA) are coloured red.

**Figure 3.**
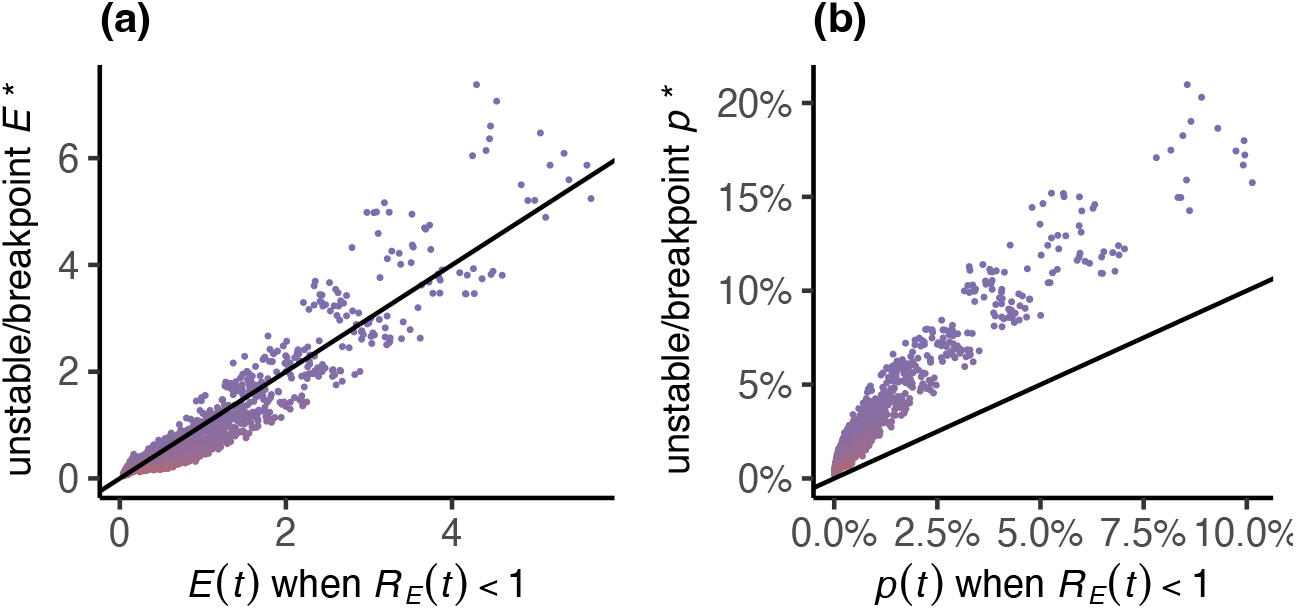
Influence of socio-ecological heterogeneity on intensity and prevalence of urogenital schistosomiasis at threshold of transmission. The mathematical transmission model was used to simulate infection dynamics sequentially from one to 25 annual rounds of mass drug administration (MDA) with praziquantel assuming a coverage of 50% and drug efficacy of 94%. The egg intensity, *E*(*t*) (eggs per 10ml of urine) and egg prevalence *p*(*t*) were recorded at the point when interruption of transmission (IoT; *R*_E_ (*t*) < 1) was achieved and are plotted against the unstable/breakpoint egg intensity (panel a) and prevalence (panel b). Variation in transmission dynamics is generated using 5,000 parameter sets from a four-dimensional Latin Hypercube sample of parameter *ω*, defining asymmetry in the human-to-human and snail-to-human components of the basic reproduction number, *R*_*H*→*S*_ and *R*_*S*→*H*_, the overdispersion of schistosomes among hosts at endemic stability, *k*^*^, and the human-to-snail population density, *N*_1_/*N*_2_. Points are coloured from purple to red for increasing *R*_0_.

### (d) Timeframes to elimination

The relationship between the number of MDA rounds required to reach IoT (Figure 4a) or elimination as a public health problem (EPHP; Figure 4b) and the endemic/stable *E*^*^ is non-linear with increasing variability for increasing *E*^*^ (there is almost no association between time to reach elimination and endemic/stable *p*^*^, Supplementary Information Figure S2). The influence of *R*_0_, *ω, k*^*^ and *N*_1_/*N*_2_ on time to IoT accords with their influence on the endemic/stable *E*^*^ but differs from the relationship with the unstable/breakpoint *E*^*^. That is, although only *R*_0_ and *k*^*^ are substantively associated with the unstable/breakpoint *E*^*^, all four variables (*R*_0_, *ω, k*^*^ and *N*_1_/*N*_2_) are associated with time to IoT. Specifically, time to IoT increases with increasing *R*_0_, and decreases with increasing *ω* (increasing asymmetry towards human-to-snail transmission), increasing *k*^*^ (i.e., *decreasing* overdispersion) and increasing *N*_1_/*N*_)_ (i.e., *decreasing* density of snail intermediate hosts, Figure 4c). Similar associations are evident with time to reach EPHP, albeit with different magnitudes (Figure 4d). Interruption of transmission is a more stringent endpoint than EPHP with consistently longer timeframes to elimination (Figure 5).

**Figure 4.**
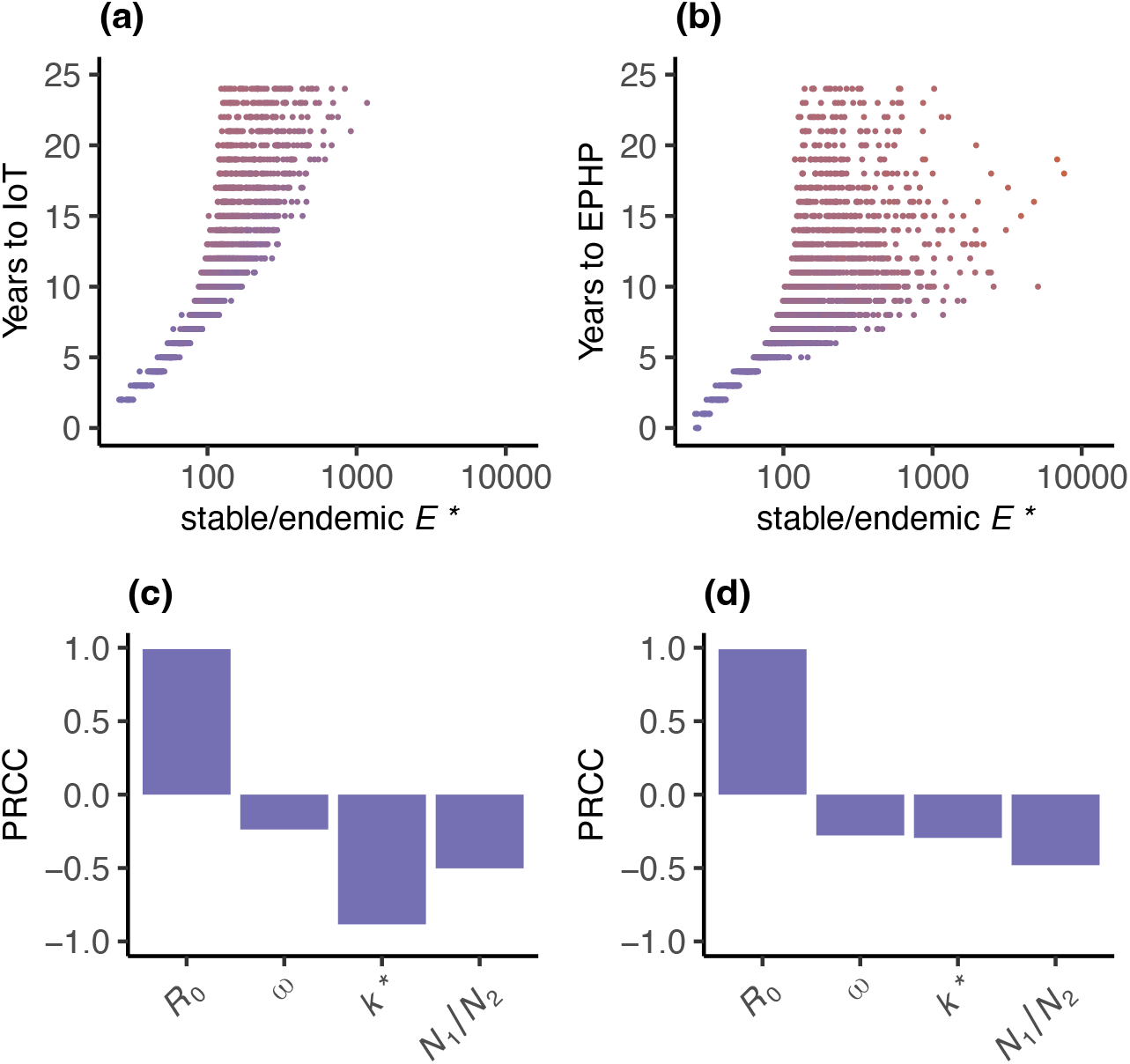
Influence of socio-ecological heterogeneity on timeframes to elimination of urogenital schistosomiasis. Variation in the stable/endemic mean egg output per host, *E*^*^ (eggs per 10 ml of urine), is generated using 5,000 parameter sets from a four dimensional Latin hypercube sample of the basic reproduction number, *R*_0_, parameter *ω*, defining asymmetry in the human-to-human and snail-to-human components of *R*_0_, *R*_*H*→*S*_ and *R*_*S*→*H*_, the overdispersion of schistosomes among hosts at endemic stability, *k*^*^, and the human-to-snail population density, *N*_1_/*N*_2_. Times to interruption of transmission (IoT) and elimination as a public health problem (EPHP) were calculated by simulating 25 annual rounds of mass drug administration (MDA) for each of the 5,000 parameter sets and determining how many years of MDA are required for the effective reproduction number to fall below 1 (*R*_E_(*t*) < 1; IoT) or the prevalence of heavy infection (< 50 eggs per 10 ml urine) to be less than 1% immediately prior to a subsequent round of MDA. Panels (a) and (b) depict the relationship between *E*^*^ and the years of MDA required to reach IoT and EPHP respectively, with colours from purple to red indicating increasing *R*_0_. Panels (c) and (d) show the influence of *R*_0_, *ω, k*^*^ and *N*_1_/*N*_2_ on times to IoT and EPHP respectively using partial rank correlation coefficients (PRCCs). The PRCC quantifies the correlation between each variable and the time to elimination while controlling for the effects of other variables.

**Figure 5.**
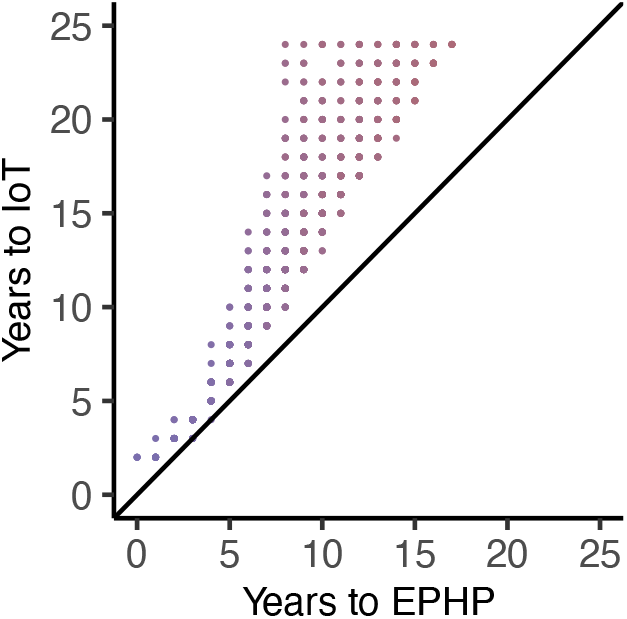
Relationship between time to interruption of transmission and elimination as a public health from for urogenital schistosomiasis under annual mass drug administration. The transmission dynamics model was used to simulate annual mass drug administration (MDA) assuming a coverage of 50% and drug efficacy of 94% for each of 5,000 parameter sets from a four dimensional Latin hypercube sample of the basic reproduction number, *R*_0_, parameter *ω*, defining asymmetry in the human-to-human and snail-to-human components of *R*_0_, *R*_*H*→*S*_ and *R*_*S*→*H*_, the overdispersion of schistosomes among hosts at endemic stability, *k*^*^, and the human-to-snail population density, *N*_1_/*N*_2_. Times to interruption of transmission (IoT) and elimination as a public health problem (EPHP) were calculated by determining how many years of MDA are required for the effective reproduction number to fall below 1 (*R*_E_(*t*) < 1; IoT) or the prevalence of heavy infection (< 50 eggs per 10 ml urine) to be less than 1% immediately prior to a subsequent round of MDA. Colours from purple to red indicate increasing *R*_0_.

## 4. Discussion

Mathematical models have become invaluable tools in the management and control of human and animal infectious disease [24, 41, 42]. For schistosomiasis and other NTDs, mathematical modelling has become increasingly used to inform high-level decision making on intervention strategies to achieve the WHO’s 2030 elimination targets [4, 5, 43]. A key, but frequently overlooked, component of this contribution is the robust and transparent communication of uncertainty [44,45]. Here, we have specifically focused on uncertainty arising from heterogeneity in socio-ecological factors that affect local transmission conditions. We show that such heterogeneity can drive substantial variation in the effectiveness of MDA and that routine epidemiological indicators of pre-intervention endemicity, such as prevalence and intensity of infection, provide limited information on parasite resilience and the likely duration of MDA required to reach elimination goals. Our findings also indicate that targeting complementary intervention efforts on reducing human exposure to infectious cercariae and reducing snail populations would facilitate reaching endpoints and accelerate time frames to elimination.

The identification of schistosomiasis hotspots in sub-Saharan Africa—despite robust implementation of MDA—has received considerable attention in the recent epidemiological literature [6-13,15] and in 2022, a provisional definition was published by the WHO [14] (but see also [20]). That pre-intervention prevalence and intensity of infection are imperfect, noisy indicators of schistosomiasis persistence is one natural explanation of these observations [15]. Socio-ecological factors driving variation in persistence and responses to MDA—such as variation in exposure and contamination, inter-individual heterogeneities and snail abundance—are typically unknown in the vast majority of affected communities. Thus, foci with ostensibly similar pre-intervention transmission conditions, as characterised by standard epidemiological indicators, may respond very differently to intervention, requiring varying durations of MDA to reach elimination endpoints. While this offers a simple explanation for the phenomenon of hotspots, it also underscores the imperative for modelling projections to include robust and realistic indicators of uncertainty in the absence of local and detailed socio-ecological data. Failing to do so may promote unrealistic expectations on the pace of progress—and ultimately the outcome—that is likely from a particular intervention strategy [44].

Where hotspots have been identified, the WHO recommends intensified biannual MDA and additional complementary strategies [14]. Exclusive reliance on MDA—even distributed to whole communities and semi-annually [2,46]—is unlikely to be sufficient to reach and sustain elimination everywhere. Integrated intervention strategies that combine MDA with improved water, sanitation and hygiene (WASH), that change behavior-particularly to reduce exposure to infectious cercariae, and that target and reduce snail populations will be essential [29,43,47]. Our results support these assertions, which also align with the historical experience of successful elimination campaigns; schistosomiasis was eradicated in Japan by aggressive targeting of snail habitats (and animal reservoirs, largely by urbanisation associated with rapid economic development) and in China, cross-sectoral integrated control (assisted by massive economic growth) has been highly effective [48]. Although the epidemiology and socioeconomic circumstances in sub-Saharan Africa are very different, there are undoubtedly lessons to be learnt from the approaches taken towards Asian schistosomiasis. A prerequisite to such multipronged interventions will be identification of communities where progress is below expectations. This will require more investment in monitoring and evaluation activities which—although emphasized by the WHO [14]—are often overlooked in the resource-scarce context of NTDs.

The WHO uses two definitions for the elimination of schistosomiasis; EPHP, defined as a prevalence of heavy infection less than 1% and IoT, defined as complete cessation of transmission or suppression to an extent such that the parasite population cannot sustain itself. The latter corresponds to intervention having suppressed the schistosome population to below the unstable equilibrium (breakpoint). Breakpoints exist because female schistosomes must mate with males to produce viable transmission stages (although the increasing overdispersion resulting from MDA means that they may become vanishing small [33]). This leads to the somewhat counter intuitive phenomenon that for dioecious macroparasites, *R*_0_ = 1 is not a threshold quantity [39]. Notwithstanding, of more practical relevance is the relationship between EPHP and IoT and specifically, whether the current 1% prevalence of heavy infection target is adequate to guard against resurgence following cessation of MDA. Although we use a highly simplified model, our results suggest that in many socio-ecological settings IoT will take considerably longer to reach than EPHP. Hence, while suppressing infection to low levels - and particularly heavy infection-has great public health benefit, it may not be a particularly useful endpoint for interventions. If interventions are halted before parasite breakpoints have been reached, resurgence is inevitable.

Similar questions have been raised about the current elimination thresholds that are used by lymphatic filariasis (LF) elimination programmes. The so-called ‘transmission assessment survey’ employs a similar prevalence threshold to determine when MDA can be stopped [49]. Yet it has been shown that IoT thresholds for LF can vary widely and that there are insufficient data to support the current threshold as a measure of sustainable transmission [50]. Ultimately, it is unpractical to suggest that elimination thresholds should be tailored specifically to individual locations for either LF, schistosomiasis or any other NTD. Resources are simply too scarce to generate the data that would be required to estimate local thresholds with an acceptable degree of precision. Rather, it must be widely recognised that endpoints based on arbitrary thresholds do not necessarily concord with IoT and that robust and agile surveillance strategies that can respond quickly and effectively to resurgent infection will be paramount in the ‘post-elimination era’ [51].

The importance of the intensity of transmission, *R*_0_, and degree of parasite overdispersion *k* on the resilience of helmith populations is well known (e.g., [23-25, 52]). The former is intuitive; the greater the potential for transmission (higher *R*_0_), the lower the parasite breakpoint. The latter arises due to the dependence of many helminths on sexual reproduction. Aggregation (overdispersion) enhances the likelihood of encountering the opposite sex within the definitive (human) host and therefore lowers the breakpoint; the more aggregated, the lower the breakpoint. However, as we have shown, the intervention effort required to reach a breakpoint for a given *R*_0_—under dynamic overdispersion resulting from MDA [33]—depends on other socio-ecological factors such as the relative importance of contamination (i.e. *R*_*H*→*S*_) versus exposure (i.e., *R*_*S*→*H*_) and the relative abundance of definitive (human) hosts compared to intermediate (snail) hosts (i.e., *N*_1_/*N*_2_). This is because of the finite abundance of snails combined with their short infectious lifespan (approximately 30 days [38]) introduces a natural restriction on the size of the parasite population. Hence, when transmission is dominated by contamination and snails are relatively scarce, the difference between the current and breakpoint population size becomes smaller, facilitating prospects of elimination. In practical terms, this explains why reducing human exposure to cercariae and controlling snail populations (i.e. increasing *ω* to decrease the relative contribution of *R*_*S*→*H*_ and decreasing *N*_2_ to increase *N*_1_/*N*_2_ in the denominator of Equation (1)) is an effective complement to MDA.

Uncertainty in mathematical models can be divided into three categories: parametric, structural and stochastic. We have here focused on parametric uncertainty relating to geographical heterogeneity in socio-ecological conditions that affect schistosomiasis transmission (i.e., potential drivers of so-called ‘biological hotspots’ as opposed to ‘operational hotspots’ relating to deficiencies in programme implementation [20]). This is not a comprehensive exploration of parametric uncertainty which extends even to parameters as fundamental as the mortality rate of adult schistosomes; a parameter that, in common with other helminths, is difficult to measure. Despite the data-scarce context of NTDs, parametric uncertainty—including that related to socio-ecological and geographic heterogeneity [25]—has received only sporadic attention [26,50,52]. This contrasts with the increasing attention on stochastic uncertainty arising through the burgeoning use of individual-based simulation models, often designed to quantify ‘elimination probabilities’ under competing intervention strategies (e.g., [5,52-55]). Such models are useful for incorporating increased programmatic realism (e.g., individual adherence patterns to MDA [56]) and, crucially, can capture stochastic effects which become more important in small populations of humans and parasites (i.e., when the latter is approaching elimination). However, the growing use of such models—which typically use fixed parameterizations—can give a false impression of precision, obfuscating uncertainty arising from underlying parameters [45]. Parametric uncertainty can be readily incorporated into stochastic (individual-based) models (e.g., as sensitivity analyses), but when the number of uncertain parameters is large, the computational cost can become substantial.

Structural uncertainty typically relates to the fundamental population processes that are incorporated into a model. For example, although acquired immunity is thought to play a substantial role in shaping the epidemiology of schistosomiasis, few models have attempted to incorporate it [57,58] partly because of limited understanding of the mechanism of action [59]. Increasingly, structural uncertainty has been addressed by using multiple models—that make fundamentally different structural assumptions—to make policy-relevant projections. This can either be achieved using so-called ensemble approaches, averaging predictions from different models [60] or expressing each as a distinct outcome associated with a particular model. The latter has largely been the preferred approach of modelling consortia such as the NTD Modelling Consortium [61,62] the Malaria Modelling Consortium [63] and the HIV Modelling Consortium [64].

Our analysis underscores the importance of robust and transparent integration of uncertainty into modelling outcomes, especially in the data-scarce context of NTDs [44]. The availability of epidemiological (prevalence and/or intensity) data on schistosomiasis is not always guaranteed (intensity data are particularly scarce) [65], and even when available will often be compatible with a range of model parameterizations (e.g., estimates of *R*_0_) and associated projections (e.g., through rounds of MDA). Longitudinal data collected during intervention can help reduce parametric uncertainty [26] but are even more scarce than baseline (pre-intervention) epidemiological indicators. In this context, it is essential that uncertainty in modelling projections (such as time frames to elimination) is expressed unambiguously both to avoid unrealistic expectations of a model’s predictive capacity and to identify when observations (such as hotspots) are truly beyond the envelope of what is expected. As expectations for the elimination of NTDs continue to rise as we move towards 2030 [1], it will be increasingly important to make explicit the limitations and uncertainty inherent to modelling projections.

## Data Availability

The model code used to produce the results from this work can be found at https://github.com/martwalker/simpleschisto

## Authors’ contributions

Conceptualization: MIN, MW; Methodology: MIN, GCM, MW; Software: MIN, MW; Formal analysis: MIN, MW; Investigation: MIN, MW; Writing – Original Draft: MIN, MW; Writing – Review and Editing; MIN, GCM, JPW, MW; Visualization: MIN, MW; Supervision: JPW, MW; Funding acquisition: JPW, MW.

## Acknowledgements

MIN acknowledges funding from a Royal Veterinary College, University of London PhD Studentship. GCM, JPW and MW were supported through the FibroScHot project which is part of the EDCTP2 programme supported by the European Union (RIA2017NIM-1842-FibroScHot). MW acknowledges funding by the Bill & Melinda Gates Foundation through the NTD Modelling Consortium (grant INV-030046).

