## Supplementary Information for "Socio-ecological heterogeneity and uncertainty in the elimination of human schistosomiasis"

### Supplementary methods

#### S1. Derivation of schistosomiasis transmission dynamics model

The human-to-snail component of the basic reproduction number, $R_{H\to S}$, is defined as the average number of infectious snails produced by a single fertile female schistosome during its lifespan in the absence of density-dependent constraints,

$$R_{H\to S} = \frac{\psi_1 N_2}{\mu_1} \qquad \text{S1}$$

Here, $\psi_1$ is a composite parameter describing the rate at which eggs are produced per fertile female schistosome per human host, the rate at which these eggs contaminate a water source (with intermediate snail hosts present), the probability that an egg develops into a miracidia and the probability that a miracidia makes effective contact with a snail to produce a patent infection (i.e. a snail shedding cercariae). Parameter $\mu_1$ is the per capita mortality rate of adult schistosomes, such that $1/\mu_1$ is the life expectancy and $N_2$ is the population density of snails.

The snail-to-human component of the basic reproduction number, $R_{S\to H}$ is defined as the average number of fertile (patent) female schistosomes produced in the human population by a single infectious snail (in the absence of density-dependent constraints),

$$R_{S\to H} = \frac{\psi_2 \rho N_1}{\mu_2} \qquad \text{S2}$$

Like $\psi_1, \psi_2$ is a composite parameter describing the rate at which cercaria are shed per infectious snail and the probability that any given cercaria makes effective contact with a human (i.e. contact leading to a patent infection). Parameter $\mu_2$ is the per capita mortality rate of infectious snails, such that $1/\mu_2$ is their life-expectancy, $N_1$ is the human host population density, and $\rho$ is the schistosome sex ratio (proportion female).

The basic reproduction number, $R_0$, is given by the product of $R_{H\to S}$ and $R_{S\to H}$ which we parameterize in terms of an asymmetry parameter, $\omega$, such that $R_{H\to S} = R_0^{\omega}$ and $R_{S\to H} = R_0^{1-\omega}$.

The dynamics in the proportion of infectious snails, $y(t)$, is given by

$$\frac{dy(t)}{dt} = \mu_1 R_0^{\omega}(N_1/N_2)W\Omega[W(t),k(t)]\Phi[W(t),k(t)][1-y(t)] - \mu_2 y(t) \quad \text{S3}$$

where $\Phi[W(t),k(t)]$ is the mating probability [1], $\Omega[W(t),k(t)]$ is a function defining the per worm density-dependent suppression of egg production [2,3], and $k(t)$ is the overdispersion parameter of a negative binomial distribution [4,5] with dynamics induced by variable adherence patterns during mass drug administration (MDA) [6]. These functions are defined in S2 Mathematical functions.

We can assume that the dynamics of infectious snails occurs on a much shorter time scale than that of adult schistosomes. Therefore, $y(t)$ in Equation S3 can be set to its equilibrium position $(y^*)$ with respect to $W(t)$,

$$y^* = \frac{\mu_1 R_0^{\omega}(N_1/N_2)W(t)\Omega[W(t),k(t)]\Phi[W(t),k(t)]}{\mu_1 R_0^{\omega}(N_1/N_2)W(t)\Omega[W(t),k(t)]\Phi[W(t),k(t)] + \mu_2} \quad \text{S4}$$

The rate of change in $W(t)$ can now be written as

$$\frac{dW(t)}{dt} = \mu_2(R_0^{1-\omega}/\rho)(N_2/N_1)y^* - \mu_1 W(t) \quad \text{S5}$$

Substituting for $y^*$ yields

$$\frac{dW(t)}{dt} = \frac{R_0^{\omega}(R_{S\to H}/\rho)\mu_1\mu_2 W(t)\Omega[W(t),k(t)]\Phi[W(t),k(t)]}{R_0^{\omega}\mu_1(N_1/N_2)W(t)\Omega[W(t),k(t)]\Phi[W(t),k(t)] + \mu_2} - \mu_1 W(t) \quad \text{S6}$$

which can be written as

$$\frac{dW(t)}{dt} = \frac{(R_0/\rho)\mu_2 W(t)\Omega[W(t),k(t)]\Phi[W(t),k(t)]}{R_0^{\omega}(N_1/N_2)W(t)\Omega[W(t),k(t)]\Phi[W(t),k(t)] + \mu_2/\mu_1} - \mu_1 W(t) \quad \text{S7}$$

as given in the main text.

#### S2. Mathematical functions

The mating probability $\Phi[W(t),k(t)]$ for monogamously mating parasites, assuming a negative binomial distribution of worms among hosts is given in [1] as

$$\Phi[W(t),k(t)] = 1 - \frac{(1-\alpha(t))^{(1+k(t))}}{2\pi}\int_{\theta=0}^{\theta=2\pi}\frac{1-\cos(\theta)}{(1+\alpha(t)\cos(\theta))^{(1+k(t))}} \quad \text{S8}$$

where $\alpha(t) = W(t)/(W(t)+k(t))$.

The function $\Omega[W(t),k(t)]$ describing the reduction in schistosome fecundity induced by density-dependent constraints is derived from the relationship between the egg output, $z$, and the number of female schistosomes per host reduction, $n$ [7]

$$z = \beta_0 n^{\beta_1} \quad \text{S9}$$

where $\beta_0$ is the maximum schistosome fecundity in the absence of density-dependent constraints and $\beta_1$ is the severity of density-dependent suppression (see main text Table 1). It follows that the reduction in fecundity, $f(n)$, expressed as a proportion of the maximum fecundity is

$$f(n) = n^{\beta_1 - 1} \quad \text{S10}$$

Assuming a negative binomial distribution of female worms among hosts, with probability that a host has $n$ female schistosomes given by,

$$P(n|W(t),k(t)) = \frac{(\rho W(t))^n}{n!} \frac{\Gamma(k(t)+n)}{\Gamma(k(t))(k(t)+\rho W(t))^n} \frac{1}{(1+\rho W(t)/k(t))^{k(t)}} \quad \text{S11}$$

it follows that the net reduction in fecundity across all schistosomes in the population, i.e., $\Omega[W(t),k(t)]$, is derived by taking the expectation of $f(n)$

$$\Omega[W(t),k(t)] = \frac{\sum_{n=1}^{n=\infty} f(n)P(n|W(t),k(t))}{\sum_{n=1}^{n=\infty} P(n|W(t),k(t))} \quad \text{S12}$$

Unlike another function forms of $f(n)$ (see for example [2,3]) there exists no closed form for Equation S12 and therefore it was calculated numerically at each time step of a model run.

The dynamics in the overdispersion parameter of the negative binomial distribution induced by MDA is modelled using the approximation derived by Collyer et al. [6] (see also [8] for an example of its implementation in a schistosomiasis transmission model). Immediately following treatment at time $\tau$,

$$k(\tau+dt) = \frac{k^*W(\tau+dt)}{(1+k^*)W^* - k^*W(\tau+dt)} \quad \text{S13}$$

where $k^*$ is the equilibrium (steady state) value of $k(t)$ and $W^*$ is the endemic/stable equilibrium worm burden. Thereafter, for $t > \tau + dt$,

$$k(t) = \frac{W(t)^2[W^* - W(\tau+dt)]^2}{\frac{{W^*}^2}{k^*}[W(t) - W(\tau+dt)]^2 + \left[\frac{W(\tau+dt)^2}{k(\tau+dt)}\right][W(t) - W^*]^2} \quad \text{S14}$$

Note that the model is parameterized using values of $k^*$ and allowing the dynamics of $k(t)$ to emerge during rounds of MDA.

#### S3. Prevalence of infection and heavy infection

The prevalence of eggs in the population $p(t)$ (i.e., the probability that a host is positive for schistosome eggs), is given by the probability that a host is infected by at least one mated female schistosome. That is,

$$\begin{aligned} p(t) &= P(n > 0|W(t),k(t))\Phi[W(t),k(t)] = \Phi[W(t),k(t)]\sum_{n=1}^{n=\infty} P(n \mid W(t),k(t)) = \Phi[W(t),k(t)] - \Phi[W(t),k(t)]P(0 \mid W(t),k(t)) \\ &= \Phi[W(t),k(t)] - \Phi[W(t),k(t)]\left(1 + \frac{\rho W(t)}{k(t)}\right)^{-k(t)} \end{aligned} \quad \text{S15}$$

The prevalence of heavy infection, $p_H(t)$—used for the definition of elimination as a public health problem (EPHP)—is calculated by determining the probability that an individual's egg output is greater than a threshold $z$,

$$p_H(t) = \Phi[W(t),k(t)]\sum_{n=(z/\beta_0)^{1/\beta_1}}^{n=\infty} P(n \mid W(t),k(t)) \quad \text{S16}$$

For *S. haematobium*, $z$ = 50 eggs per 10ml urine as per the World Health Organization [9] definition of heavy infection (see Table 1 main text). Note that $p(t)$ and $p_H(t)$ and somewhat hypothetical, assuming a perfect diagnostic (100% sensitivity; 100% specificity) to detect egg output.

#### S4. Partial rank correlation coefficients

Parameters $R_0, \omega, N_1/N_2$ and $k^*$ were sampled from a four-dimensional Latin hypercube, with each parameter assigned a uniform distribution (see Table 1 for ranges). Partial rank correlation coefficients (PRCCs) were used to quantify the influence of these parameters on the model's stable and unstable/breakpoint equilibria and the number of rounds of annual MDA (assuming a nominal 50% coverage and praziquantel efficacy of 94%) required to reach EPHP or interruption of transmission (IoT) (see main text for details). The PRCC quantifies the relationship between a rank-transformed input variable $x_j$ (i.e., $R_0, \omega$, $N_1/N_2$ and $k$ ) and a rank-transformed model output $y$ (i.e., an equilibrium value or number of MDA rounds to reach EPHP or IoT) having discounted the effects of other inputs [10]. The PRCC is the correlation coefficient between the residuals $(x_j - \hat{x}_j)$ and $(y - \hat{y})$, where $\hat{x}_j$ and $\hat{y}$ are derived from the linear regression models,

$$\hat{x}_j = c_0 + \sum_{\substack{p=1 \\ p \neq j}}^{k} c_p x_p \qquad \text{S17}$$

and

$$\hat{y} = b_0 + \sum_{\substack{p=1 \\ p \neq j}}^{k} b_p x_p \qquad \text{S18}$$

where $b_0$ and $c_0$ are intercept terms and $b_p$ and $c_p$ are regression coefficients. The PRCC, $r_{x_j y}$, is then given by

$$r_{x_j y} = \frac{\sum \left((x_j - \hat{x}_j) - E[(x_j - \hat{x}_j)]\right)((y - \hat{y}) - E[(y - \hat{y})])}{\sum \left((x_j - \hat{x}_j) - E[(x_j - \hat{x}_j)]\right)^2 \sum \ ((y - \hat{y}) - E[(y - \hat{y})])^2} \qquad \text{S19}$$

where $E[\cdot]$ denotes the sample mean and, for brevity, explicit summation over each individual parameter values and model outputs are suppressed.

### Supplementary figures

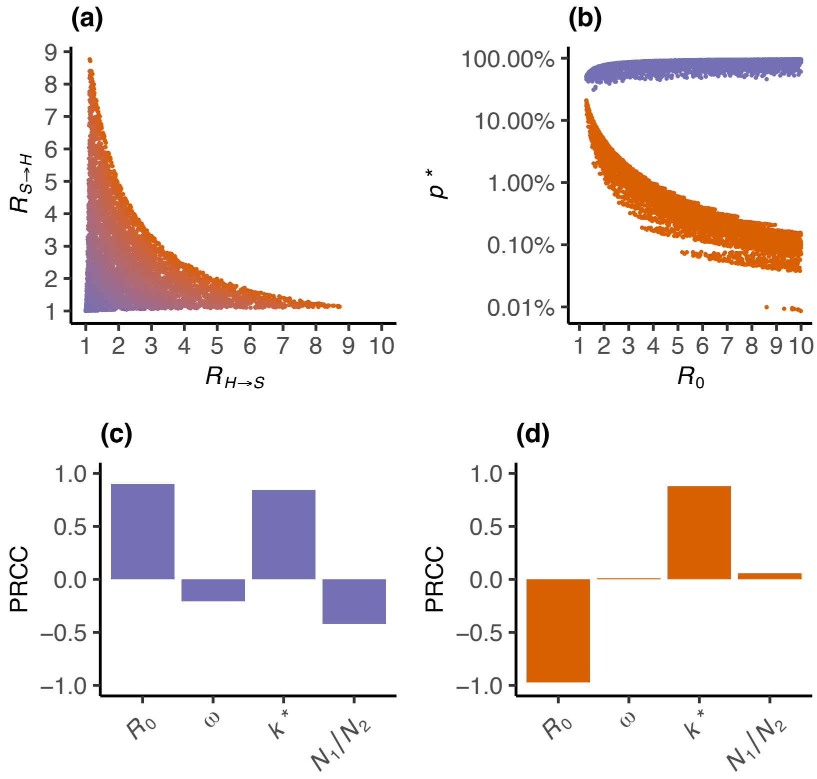


**Figure S1**. Influence of socio-ecological heterogeneity on stable/endemic and unstable/breakpoint *Schistosoma haematobium* infection prevalence. Variation in the stable/endemic (purple points and bars) and unstable/breakpoint (red points and bars) equilibrium mean prevalence, $p^*$, is generated using 50,000 parameter sets from a four dimensional Latin hypercube sample of the basic reproduction number, $R_0$, parameter $\omega$, defining asymmetry in the human-to-human and snail-to-human components of $R_0$, $R_{H\to S}$ and $R_{S\to H}$, the overdispersion of schistosomes among hosts at endemic stability, $k^*$, and the human-to-snail population density, $N_1/N_2$. Panel (a) shows the relationship between $R_{H\to S}$ and $R_{S\to H}$ arising from sampling $\omega \in (0,1)$ and $R_0 \in (1,10)$, with colours from purple to red indicating increasing $R_0$. Panel (b) shows the relationship between the stable/endemic (purple) and unstable/breakpoint (red) $E^*$ and $R_0$, with variation driven by parameters $\omega$, $k^*$ and $N_1/N_2$. Panels (c) and (d) show the influence of $R_0$, $\omega$, $k^*$ and $N_1/N_2$ on the stable/endemic $p^*$ (purple bars) and the unstable/breakpoint $p^*$ (red bars) using partial rank correlation coefficients (PRCCs). The PRCC quantifies the correlation between each variable and $E^*$, while controlling for the effects of other variables.

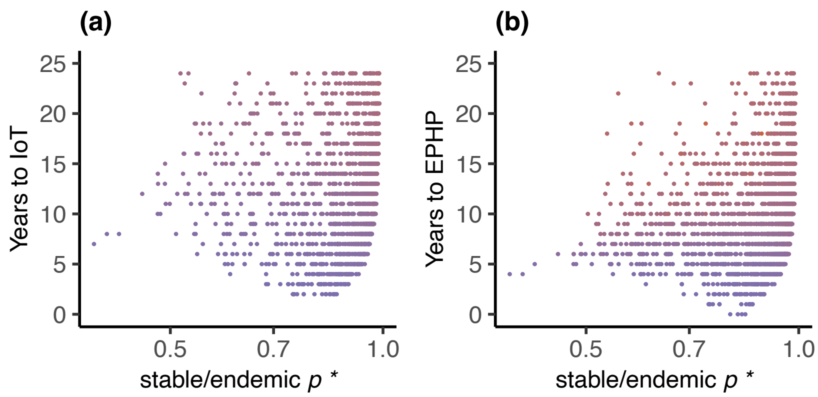


**Figure S2.** Relationship between stable/endemic prevalence or urogenital schistosomiasis and timeframes to elimination. Variation in the stable/endemic prevalence, $p^*$, is generated using 5,000 parameter sets from a four dimensional Latin hypercube sample of the basic reproduction number, $R_0$, parameter $\omega$, defining asymmetry in the human-to-human and snail-to-human components of $R_0$, $R_{H\to S}$ and $R_{S\to H}$, the overdispersion of schistosomes among hosts at endemic stability, $k^*$, and the human-to-snail population density, $N_1/N_2$. Times to interruption of transmission (IoT) and elimination as a public health problem (EPHP) were calculated by simulating 25 annual rounds of mass drug administration (MDA) for each of the 5,000 parameter sets and determining how many years of MDA are required for the effective reproduction number to fall below 1 ($R_E(t) < 1$; IoT) or the prevalence of heavy infection (< 50 eggs per 10 ml urine) to be less than 1% immediately prior to a subsequent round of MDA. Panels (a) and (b) depict the relationship between $p^*$ and the years of MDA required to reach IoT and EPHP respectively, with colours from purple to red indicating increasing $R_0$.
